# Real-time impacts of air pollution on the health, well-being, and daily life of children and young people in Delhi and Dhaka

**DOI:** 10.1101/2025.10.14.25338037

**Authors:** Constance Bwire, Gabrielle Bonnet, Ana Bonell, Rachel Juel, James Milner, Shunmay Yeung, Robert Hughes

## Abstract

Air pollution is a major global health threat, with children and young people (CYP) among the most vulnerable. Delhi (India) and Dhaka (Bangladesh) are two of the world’s most polluted cities, with persistently high levels of fine particulate matter (PM₂.₅). This study aimed to generate CYP-centered evidence on the real-time impacts of air pollution in these cities by comparing health, well-being, and daily activities during periods of high air pollution and good air quality, while also capturing CYP’s ideas for air quality management. A cross-sectional, real-time digital survey was conducted in Delhi on January 9-10 and 15 March 2025, and in Dhaka on January 21-22 and March 13-16, 2025. For both cities, the January dates correspond to a period with high air pollution (PM₂.₅ > 125.5 µg/m³) and the March dates to good air quality (PM₂.₅ ≤ 35.4 µg/m³). Participants included CYP aged 13–29 years and parents of children under 18. Recruitment was carried out online. Data on health symptoms, well-being (general feelings and sleep quality), and daily activity disruptions were analyzed using descriptive statistics, chi-square tests, and regression models adjusted for demographics. Responses to open-ended questions were thematically coded. A total of 814 eligible responses were collected (Delhi = 365; Dhaka = 449). High-pollution days were associated with significantly higher reports of itchy eyes, respiratory difficulties, headaches, skin irritation or rash, diarrhoea or vomiting, low mood, anxiety or stress, and difficulty concentrating. These associations remained significant after adjusting for demographics. Disruptions to daily activities also increased, including reduced physical activity and greater odds of being late or missing school or work, meetings, social events, and healthcare, as well as a greater need for family assistance (adjusted odds ratios approximately 3.8 to 4.8). In Delhi, changes were more pronounced across most outcomes, particularly a sharper drop in physical activity. In Dhaka, the same pattern was observed, along with additional increases in sore throat, cough, food insecurity, and difficulty accessing clean water. Participants’ suggestions clustered around five themes: cleaner environments, stronger communities, improved healthcare and education, pollution and technology solutions, and other ideas. High air pollution was linked to widespread impacts on health, well-being, and daily routines among CYP. Their proposed solutions offer insights for participatory and equitable approaches to urban air quality management.

## Background

Air pollution is a leading global health threat, associated with an estimated 6.7 million premature deaths each year [1,2]. According to the World Health Organization, 99% of the global population breathes air that exceeds the recommended health-based limits [1,3]. Urban areas are particularly affected, with traffic emissions, industrial activity, construction dust, and open waste burning all contributing to dangerously high levels of pollution [1,4]. The rapid pace of urbanization in low- and middle-income countries (LMICs) intensifies these challenges, straining health systems and deepening existing social inequalities [4]. Climate change compounds these pressures by contributing to more frequent and severe pollution episodes through altered atmospheric conditions, increased wildfire activity, rising temperatures which further exacerbate the conditions for poor air quality and wildfires [5].

Among air pollutants, fine particulate matter (PM₂.₅), particles with a diameter of 2.5 micrometres or smaller, pose the greatest threat to health [2]. Owing to their microscopic size, PM₂.₅ particles can penetrate deep into the lungs and bloodstream, exacerbating a range of health conditions, including cardiovascular and respiratory diseases; neuroinflammation; and mental health disorders such as anxiety, depression, and cognitive decline [1,2,6].

Children and young people (CYP) are among the most vulnerable to the health effects of PM₂.₅. The development of their respiratory and immune systems, higher respiratory rates, and smaller airways increase both exposure and physiological susceptibility (6,7). Many spend extended periods outdoors and lack the autonomy to protect themselves from such pollutants. Early-life exposure to PM₂.₅ can impair lung development, contribute to long-term respiratory and cardiovascular conditions, and increase the risk of chronic disease across the life course (2). In addition to physical health, PM₂.₅ pollution has been shown to negatively affect CYP’s mental well-being, sleep quality, school attendance, and emotional development (7,8). These findings suggest that PM₂.₅ not only undermines physical health but also disrupts daily routines, emotional well-being, and long-term life opportunities.

The risks associated with exposure to PM₂.₅ are especially pronounced in South Asia, which is home to many of the world’s most polluted cities [7]. Delhi, India, and Dhaka, Bangladesh, consistently rank among the worst globally for annual PM₂.₅ concentrations, with an annual average of approximately 90 µg/m³, which is almost twenty times the WHO’s recommended limit of 5 µg/m³ [1,7]. This persistent pollution is fuelled by dense urban populations, widespread industrial emissions, heavy traffic, and solid fuel use [4]. For residents of informal settlements, exposure is further compounded by inadequate housing, poor ventilation, and limited access to health services or protective infrastructure [4,8]. These intersecting structural conditions place CYP at particularly high risk, creating a dual burden of indoor and outdoor exposure with few available coping mechanisms. Delhi and Dhaka exemplified the convergence of rapid urban growth, environmental degradation, and social inequality, which characterize much of urban South Asia and the rest of the Global South.

While evidence on air pollution and health is extensive and links exposure to respiratory, cardiovascular, neurological, and emotional outcomes (5,7,9), gaps in how these risks are studied and understood remain. Much of the literature relies on retrospective data collection, clinical data, or environmental monitoring [9–13]. While these approaches are vital for identifying air pollution trends and overall health impacts, they often involve significant delays between the occurrence of pollution periods and data collection, creating delays and recall biases that obscure the short-term impacts and lived experiences of PM₂.₅ exposure. As a result, the immediate effects on daily routines, emotional health, and short-term well-being are often poorly understood.

The absence of real-time, CYP-centred evidence is particularly concerning in rapidly urbanizing South Asian cities. Pollution periods occur frequently, yet institutional responses are often slow, fragmented, or absent. Without timely, context-specific insights, policymaking risks being reactive and exclusionary, failing to account for the lived experiences of those most directly affected. Understanding how CYP themselves experience high-pollution episodes and envision improvements to be made is critical not only for capturing the full scope of health impacts but also for designing interventions that are inclusive, equitable, and responsive to local realities.

This study addresses these gaps by exploring how CYP in Delhi and Dhaka experienced periods of high air pollution, identified in real time through air quality monitoring. Using a digital rapid-response survey design, data were collected during verified high-pollution periods (daily average PM₂.₅ >125.5 µg/m³) and repeated during periods of good air quality (daily average PM₂.₅ ≤ 35.4 µg/m³ for comparison. The digital surveys captured self-reported physical symptoms, emotional states, disruptions to daily routines, and perceptions of city-level responses. Open-ended questions invite participants to propose solutions, offering a CYP perspective into the evidence base.

By linking real-time PM₂.₅ monitoring with rapidly deployed digital surveys, this study aimed to generate CYP-centered evidence on the impacts of air pollution in Delhi and Dhaka. Specifically, it examined differences in general well-being, health symptoms, and disruptions to daily activities between periods of high and good air quality while also capturing CYP’s suggestions for improving city-level sustainability. This approach provides a novel methodology for capturing the immediate, subjective impacts of air pollution events as they unfold, moving beyond retrospective assessments to reflect real-time health and daily activity disruptions. As PM₂.₅ pollution intensifies globally, cities must respond not only to rising health burdens but also to the lived experiences and ideas of those most affected. This study contributes actionable, CYP- informed evidence for shaping responsive, sustainable, and equitable environmental health policies in South Asia and other rapidly urbanizing regions worldwide.

## Methods

### Study design

This cross-sectional study aimed to examine the real-time impacts of air pollution on the health, well-being (general feelings and sleep quality), and daily life of CYP in two major South Asian cities: Delhi (India) and Dhaka (Bangladesh). Data were collected during periods of high air pollution, as well as during good air quality, allowing for comparisons to be made between cities and across pollution periods. Data were collected in Delhi during high air pollution on Thursday to Friday, 9-10 January 2025, and during good air quality on Saturday, 15 March 2025; and in Dhaka during high air pollution on Tuesday to Wednesday, 21–22 January 2025, and during good air quality on Thursday to Sunday, 13-16 March 2025. This study extends previous research on climate-related extreme periods and the health of CYP [14,15]

### Study area

Delhi (India) and Dhaka (Bangladesh) were selected from a larger group of 178 cities identified by the Children, Cities and Climate (CCC) Action Lab. The CCC Action Lab, led by the London School of Hygiene & Tropical Medicine (LSHTM), works to amplify the voices of CYP and support equitable climate solutions in fast-growing cities. These 178 cities were chosen for their large CYP populations, rapid urban development, and existing links with CCC partners (S1 Text). Delhi and Dhaka were included in this study because high levels of air pollution were recorded during the study period. An automated system tracking air quality helped identify these periods, allowing the research team to launch the survey quickly and collect timely data.

Delhi is India’s capital and second most populous metropolitan area, with over 30 million residents [16]. It faces recurrent air quality crises driven by vehicular emissions, industrial activity, construction dust, solid fuel use in peri-urban areas, and seasonal crop residue burning [4]. Meteorological conditions, particularly winter temperature inversions, trap pollutants close to the ground and contribute to prolonged smog periods [17]. Rising surface temperatures in cities have also been linked to the “heat‒pollution paradox”, in which warming exacerbates pollution hotspots and increases local exposure risk [18].

Dhaka, the capital of Bangladesh, is one of the fastest-growing megacities in the world, home to more than 20 million people [16]. Its air pollution is driven by heavy traffic congestion, unregulated brick kilns, industrial discharges, biomass burning, and rapid construction [4]. The city has experienced a marked increase in daytime urban heat island (UHI) intensity, rising from 2.20 °C in 2000 to 3.18 °C in 2019, largely due to rapid urbanization and declining green cover [19]. This warming hinders pollutant dispersion, increases cooling demand, and increases exposure risk. Extended hot, dry spells further intensify pollution, with annual PM₂.₅ levels reaching nearly 20 times the WHO guidelines [20].

Across South Asia, unplanned urban expansion and limited green infrastructure have compounded the effects of climate variability, creating persistent “brown clouds” of air pollution that overlap with rising heat risks [21,22]. Both Delhi and Dhaka exemplify how climate change, rapid urbanization, and weak regulatory enforcement converge to create sustained periods of poor air quality, disproportionately affecting CYP, particularly those with inadequate housing, poor ventilation, and limited access to health services [4,22,23].

### Air Quality Monitoring and Participant Recruitment

The air quality in both cities was monitored continuously via a linked automated system using publicly available air quality data. Participant recruitment was carried out in two defined air quality periods: high-pollution periods and good air quality periods. The identification of air quality status was based on real-time and historical PM₂.₅ data, accessed through the IQAir (AirVisual) application programming interface (API) (https://www.iqair.com/world-air-quality-api). A high air pollution period was defined as 24 or more consecutive hours during which PM₂.₅ concentrations exceeded 125.5 µg/m³, aligning with the "very unhealthy" category of the U.S. Environmental Protection Agency (EPA) Air Quality Index (AQI) [24,25]. By contrast, good air quality was defined as a 24-hour mean PM₂.₅ ≤ 35.4 µg/m³. This corresponds to the AQI Moderate category (AQI ≤ 100), a level that does not typically trigger public health advisories in India or Bangladesh [26]. This threshold is also below national 24-hour PM₂.₅ standards i.e., 60 µg/m³ in India and 65 µg/m³ in Bangladesh, thereby identifying days that are cleaner than usual in Delhi and Dhaka [20,27]. An automated monitoring system was developed via Airtable [15], which continuously tracked PM₂.₅ data and triggered alerts when the criteria for high air pollution were met.

In both air quality periods, participants were recruited through paid digital advertisements on Meta (Facebook and Instagram) and Google platforms (including YouTube, Gmail, and Google Search). Advertisements were targeted at two key populations: young people aged 13-29 years and adults aged 18 years and older who were parents or guardians of children under 18 years [15]. To be eligible, individuals needed to reside in the selected city, have internet access, and be able to complete an online survey. Participation was voluntary and anonymous, and no identifying personal data was collected.

### Data collection and management

Data was collected through a self-administered online survey hosted on Typeform (https://www.typeform.com), which is designed to be mobile-friendly and accessible across digital devices. Recruitment was carried out via targeted digital advertisements on Meta (Facebook and Instagram) and Google platforms (including YouTube, Gmail, and Google Search), with a focus on the residents of Delhi and Dhaka. Survey windows were: Delhi, 9–10 Jan 2025 (48 hours, high air pollution) and 15 Mar 2025 (24 hours, good air quality); Dhaka, 21–22 Jan 2025 (48 hours high air pollution) and 13–16 Mar 2025 (96 hours, good air quality) (see S1 File). Surveys were open only during these windows to capture real-time responses for each city. In line with the event definitions and windows, data collection used a pragmatic, event-triggered online design without an a priori power calculation; sample size was set by the exposure periods.

The survey took approximately five minutes to complete and included a combination of closed- and open-ended questions. The survey covered demographic factors, well-being, air-pollution-related health symptoms, daily activities, perceptions of city response, and air quality improvement ideas. Well-being was captured with both general feelings and sleep quality, each on 5-point scales (very bad to very good). Health symptoms and daily activity disruptions were recorded as binary (yes/no) items (e.g., itchy eyes or respiratory difficulties; being late or missing school/work or missed healthcare appointments). Physical activity used ordered categories, from none to ≥60 minutes /day. Perceptions included concern about high air pollution (0–10; higher = greater concern) and satisfaction with community preparedness/response (5-point ordered scales (very satisfied to very unsatisfied). Open-ended item: participants provided free-text suggestions to improve their city’s air quality; responses were thematically coded. No personally identifiable information was collected (See S2 Text and S3 Text).

The survey was prepared in the predominant language of each city, such as Hindi for Delhi (S2 Text) and Bengali for Dhaka (S3 Text). Draft translations were first generated via generative artificial intelligence (AI) tools (e.g., ChatGPT) and subsequently reviewed and refined by native speakers with knowledge of local dialects and terminology.

Survey responses were collected through Typeform, hosted on Amazon Web Services, which complies with the General Data Protection Regulation (GDPR) and is certified under ISO/IEC 27001. To enable real-time monitoring of participation and data quality during both high- and low-pollution periods, responses were temporarily synced to encrypted Google Sheets accessible only to authorized researchers via institutional accounts with two-factor authentication. The final datasets were stored on password-protected, encrypted devices. No personal identifiers, IP addresses, or tracking data were collected, ensuring complete anonymity and confidentiality. This study was approved by the LSHTM’s Research Ethics Committee (Reference 31044). Participants provided electronic informed consent via a mandatory checkbox at the start of the Typeform survey after reading the online information sheet [28]. Consent selections were automatically recorded in the response record, and participants could not proceed without consenting. For participants <18 years, a parent provided e-consent and completed the survey on the child’s behalf. Witnessing was not required by the ethics approval for this electronic procedure; no signed paper forms were used [28]. To promote transparency and local engagement, the offices of the mayors in Delhi and Dhaka were informed of the study before data collection (see S4 Text for the sample email) [15].

### Data analysis

All survey responses were checked and cleaned prior to analysis. Entries that were incomplete or did not meet eligibility requirements were excluded, and duplicate submissions were removed via the duplicate case function in the Statistical Package for the Social Sciences (SPSS). Most questions in the survey were compulsory, which reduced the amount of missing data. Responses such as ‘I don’t know’ and ‘Prefer not to say’ were retained and treated as valid categories.

To allow cross-city thematic analysis, the responses were translated into English. Draft translations were generated with AI-assisted tools and then reviewed and refined by bilingual speakers fluent in Hindi, Bengali, and English to ensure accuracy and cultural relevance.

Data collected during both high-pollution and good air quality periods were analyzed to examine variations in health, well-being, daily routines, and participant experiences. Quantitative analyses were conducted via the IBM Statistical Package for the Social Sciences (SPSS) Statistics Version 30, whereas qualitative responses were explored thematically via the NVivo qualitative analysis software Version 15. Descriptive statistics summarized demographic profiles, self-reported health, and disruptions to daily life. The analyses were stratified by air quality condition (high air pollution vs. good air quality). Pearson’s chi-square tests were applied to assess group differences in demographics and outcomes, including sex, reported symptoms, and disruptions to activities.

Associations between air quality periods and health outcomes were examined via binary logistic regression models. Ordinal variables such as self-rated health and sleep quality were analyzed via ordinal logistic regression, with scores ranging from 1 (very poor) to 5 (very good). The models included air quality and demographic factors (age group, sex, income, and city). Binary outcomes (e.g., presence of specific symptoms, missed school or work) were also assessed through logistic regression with adjustment for confounders.

For the open-ended questions on improving urban health and sustainability, the responses were analyzed thematically via a structured five-step previously published methodology [15]. This process began with repeated reading for familiarization, followed by coding with a combination of inductive and in vivo strategies. Codes were grouped into broader themes, which were checked for consistency via NVivo’s coding comparison tool. Themes were refined and illustrated with participant quotations, enabling the identification of shared priorities and lived experiences across both cities.

## Results

### Survey participation and demographics

A total of 814 eligible survey responses were collected across the two cities. These included 365 from Delhi and 449 from Dhaka, during periods of good air quality (n = 272) and high air pollution (n = 542) (Fig 1). In Delhi, episodes of high air pollution were recorded on 9 and 10 January 2025, with 24 hour average PM₂.₅ concentrations of 182 µg/m³ and 171 µg/m³, respectively. These levels corresponded to AQI values classified as “Hazardous” (9 January) and *“*Very Unhealthy*”* (10 January) under the U.S. EPA system [24,25]. According to India’s National Air Quality Index (NAQI), both days were categorized as *“*Very Poor”[27]. A good air quality day was detected on 15 March 2025, with a 24 hour average PM₂.₅ concentration of approximately 35.4 µg/m³, placing it at the upper threshold of the *“*Moderate*”* category according to the U.S. AQI [24,25]. Under India’s NAQI, this day was classified as *“*Satisfactory*”* (Central Pollution Control Board., 2024).

**Fig 1:**
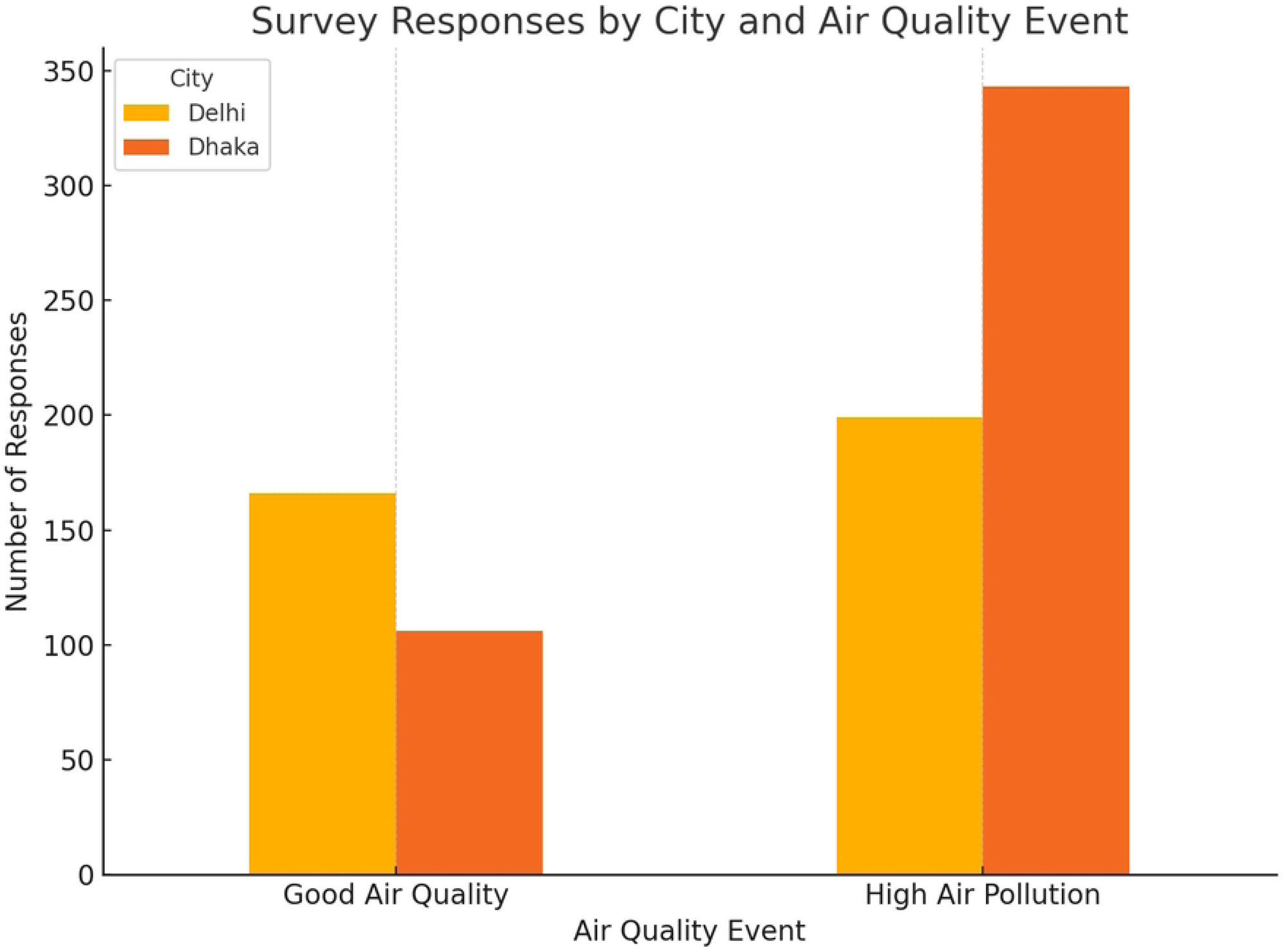
Survey participation across cities and air quality periods. The figure shows the number of eligible participants from Delhi and Dhaka during periods of high air pollution and good air quality.

In Dhaka, a high-pollution episode occurred on 21–22 January 2025, with 24 hour average PM₂.₅ concentrations of 118 µg/m³ and 117 µg/m³, both categorized as “Unhealthy*”* according to the U.S. EPA AQI and Bangladesh’s national AQI system [25]. A period of relatively improved air quality was recorded between 13 and 16 March 2025, during which 24 hour average PM₂.₅ levels ranged from 30 to 33 µg/m³, consistent with the *“*Moderate” category according to Bangladesh’s national AQI system and the U.S. EPA categories [25,29].

The number of participants was greater during high-pollution periods, particularly in Dhaka (n = 343) (Fig 1).

Among all participants, 52.8% (n = 430) were parents. In Delhi, the proportion of parents was similar across periods, i.e., 34.6% during good air and 35.7% during high air pollution (S1 Table). In Dhaka, parent participation was higher during high air pollution (77.0%) than during good air quality. The overall gender distribution was 61.3% (n = 499) male, 34.2% (n = 278) female, and 4.5% (n = 37) other/prefer not to say. Gender-city comparisons revealed statistically significant differences by air quality period in both Delhi (p < 0.001), where there were more females in high air pollution periods, and Dhaka (p < 0.001), where there were more males in high air pollution periods; however, these differences were not statistically significant at the aggregate level (p = 0.354).

In Delhi (high air pollution, n=199; good air quality, n=166), during good air quality periods, participants reported mid-range incomes more frequently: $100–$499 (10.2% vs 4.5%), $500–$1,499 (6.0% vs 1.5%), and $1,500–$4,000 (18.7% vs 2.5%). During high air pollution, participants had higher proportions at the very low end (<$100: 21.1% vs 18.1%) and the upper end (≥$4,000: 6.0% vs 2.4%), and a larger share responded “I don’t know” (64.3% vs 44.6%). In Dhaka (high air pollution, n=343; good air quality, n=106), during good air quality participants were concentrated in the lowest bracket (<$100: 69.8% vs 32.7%), whereas the high-pollution group more often reported $100–$499 (25.1% vs 3.8%) and selected “I don’t know” more frequently (33.5% vs 18.9%); other brackets were small in both groups (≤4.4%) (S1 Table).

The sample was broadly distributed across age groups; the largest groups were 10–14 years (22.1%, n = 180) and 15–19 years (20.8%, n = 169). In Delhi, the most represented age group was 10–14 years (25.2%, n = 92/365), and the distribution did not differ by period (p = 0.122). In Dhaka, age-group distributions differed by period (p < 0.001), with 15–19-year-olds more frequent during high air pollution (28.3%, n = 97/343) than during good air (16.0%, n = 17/106), 25.4% (n = 114/449) overall across periods. Among parent participants, children’s age groups differed by period in Dhaka (p < 0.001). The largest group was pre-teens aged 10–14 years (33.8%, n = 98/290), comprising a greater share during high air pollution (39.1%, n = 93/238) than during good air (9.6%, n = 5/52). In Delhi, the distribution did not differ by period (p = 0.309). A summary of key demographics is shown in Fig 2, with full counts in S1 Table.

**Fig 2:**
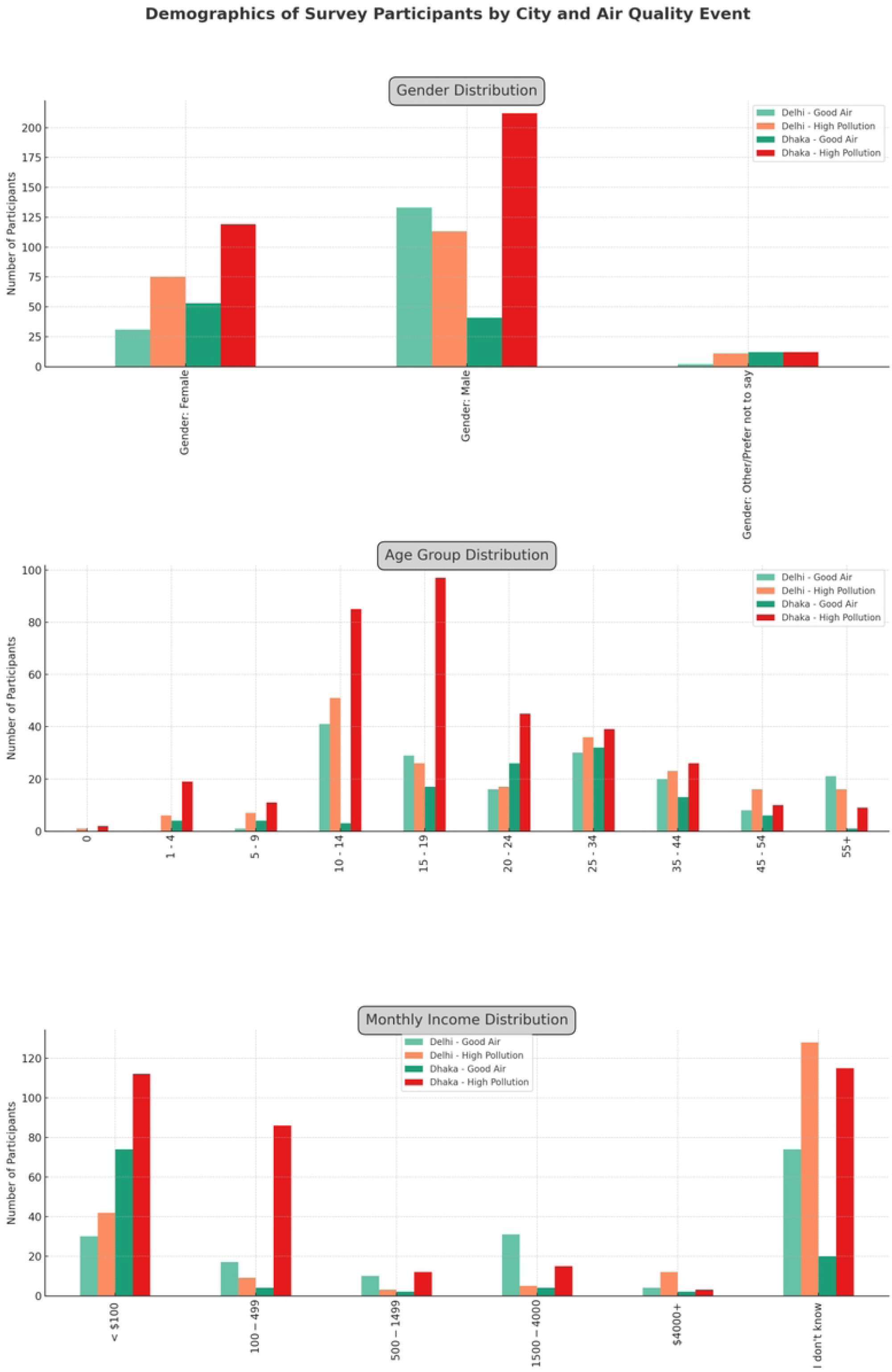
Demographics of survey participants by city and air quality period. The key demographic distribution of survey participants from Delhi and Dhaka during periods of good air quality and high air pollution, showing variations in gender, age groups, and monthly income across the two cities and air quality periods.

### General well-being

Self-reported feelings differed by air-quality period in Delhi, Dhaka, and the combined sample (all p < 0.001). In Delhi, during high air pollution, n = 46/199 (23.1%) reported “Bad” and n = 66/199 (33.2%) reported “Good,” whereas during the good air quality period, n = 4/166 (2.4%) reported “Bad” and n = 28/166 (16.9%) reported “Good.” Reports of “Ok” and “Very Good” declined from n = 56/166 (33.7%) and n = 75/166 (45.2%) under good air to n = 12/199 (6.0%) and n = 74/199 (37.2%) during high air pollution, respectively; “Very Bad” was rare (n = 4/365 across periods). In Dhaka, feelings also shifted with air quality: during high air pollution n = 89/343 (26.0%) reported “Ok” and n = 152/343 (44.3%) reported “Very Good,” whereas during the good- air period both “Ok” and “Very Good” were n = 15/106 (14.2%) (with “Very Bad” n = 9/449 across periods). When pooling cities (N = 814), positive feelings (“Good/Very Good”) dominated during good air (n = 186/272 (68.4%)), while negative feelings (“Bad/Very Bad”) were n = 15/272 (5.5%) and “Ok” n = 71/272 (26.1%). Compared to periods of good air quality, in periods of high air pollution, negative feelings more than doubled (n = 65/542 (12.0%)), “Ok” decreased (n = 101/542 (18.6%)), and positives were n = 376/542 (69.4%). City-level and combined distributions appear in S1 Fig and Fig 3; full cross-tabulations are provided in S2 Table.

**Fig 3:**
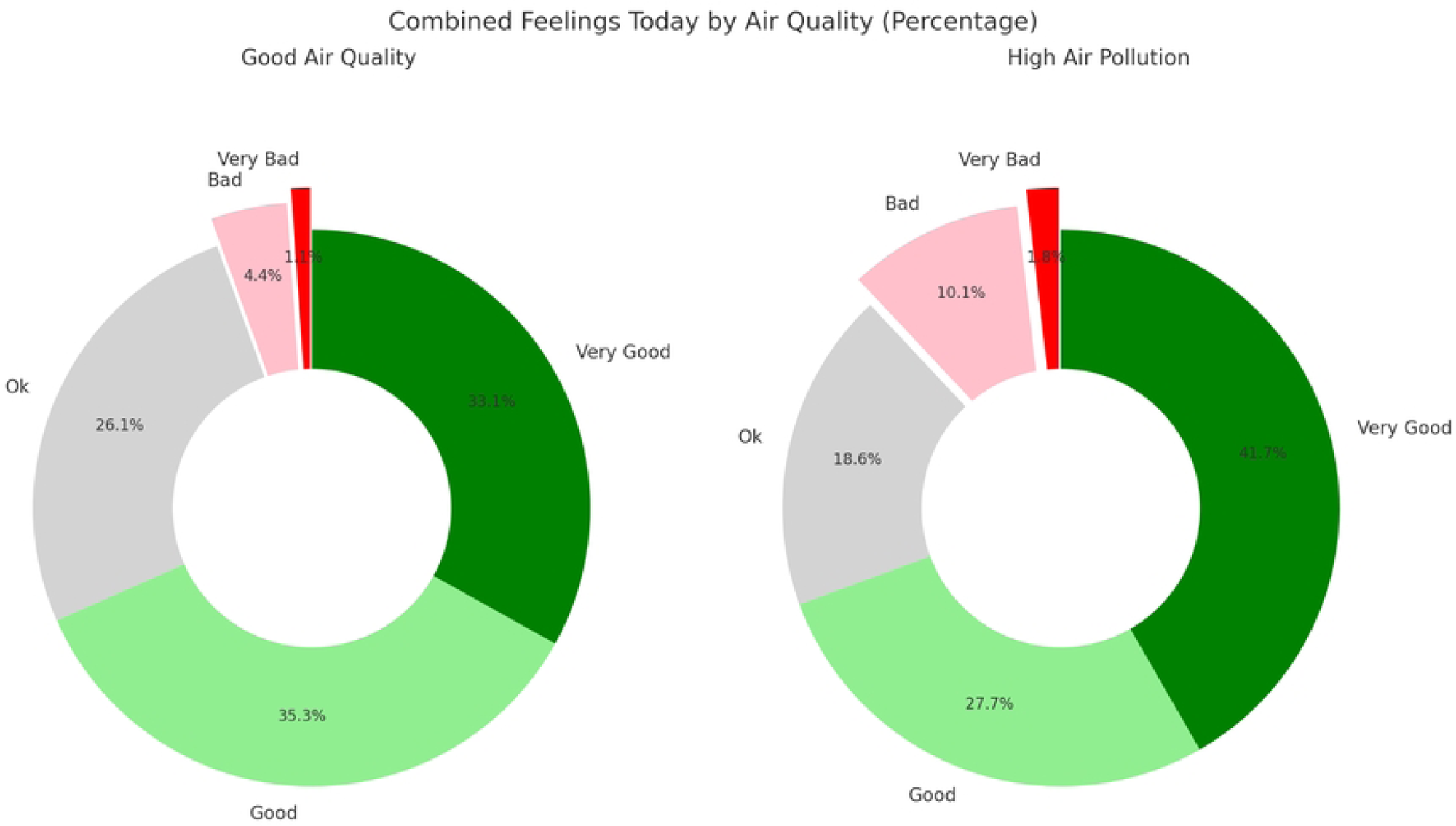
Percentage of reported general feelings in Delhi and Dhaka combined during good air quality and high air pollution periods. Distribution of participants’ self-reported feelings across varying air quality periods.

When the data from both cities were combined (N = 814), positive general feelings (good and very good) dominated during periods of good air quality, accounting for 68.4% of the responses, whereas negative feelings (bad and very bad) were reported by only 5.5% of the participants; the remaining 26.1% reported neutral feelings (“Ok”). Under high air pollution, the percentage of negative feelings more than doubled to 12.0%, while the percentage of positive feelings slightly increased to 69.4%, and the percentage of neutral responses decreased to 18.6%.

These differences were statistically significant in both cities (p < 0.001) and at the total sample level (p < 0.001), indicating a clear relationship between air quality and self-reported general feelings. The combined self-reported feelings for Delhi and Dhaka are presented in Fig 3, while detailed distributions are provided in S1 Fig and S2 Table.

Ordinal logistic regression was conducted following chi-square analyses to examine the associations between air quality periods (main predictor), participants’ demographic characteristics (covariates), and general well-being variables, i.e., sleep quality and general feelings (outcomes). While chi-square analyses revealed significant differences across air quality periods, ordinal regression indicated that air quality periods were not a significant predictor of either outcome (p > 0.05). Full estimates are provided in S2 Table.

Sleep quality changes with air quality. In Delhi (p < 0.001), most participants reported “Ok” sleep during good air quality (39.8%, n = 66), but this shifted to “Good” sleep during high air pollution (32.7%, n = 65). In Dhaka (p < 0.001), “good” sleep was most common under good air quality (67.0%, n = 71), whereas during high air pollution, the majority reported either “good” (33.8%, n = 116) or “very good” sleep (32.4%, n = 111). When data from both cities were combined (N = 814), “good” sleep was the most common response during both good air quality (45.2%, n = 123) and high air pollution (33.4%, n = 181) periods. These differences were statistically significant (p < 0.001) (Fig 4 & S2 Table).

**Fig 4:**
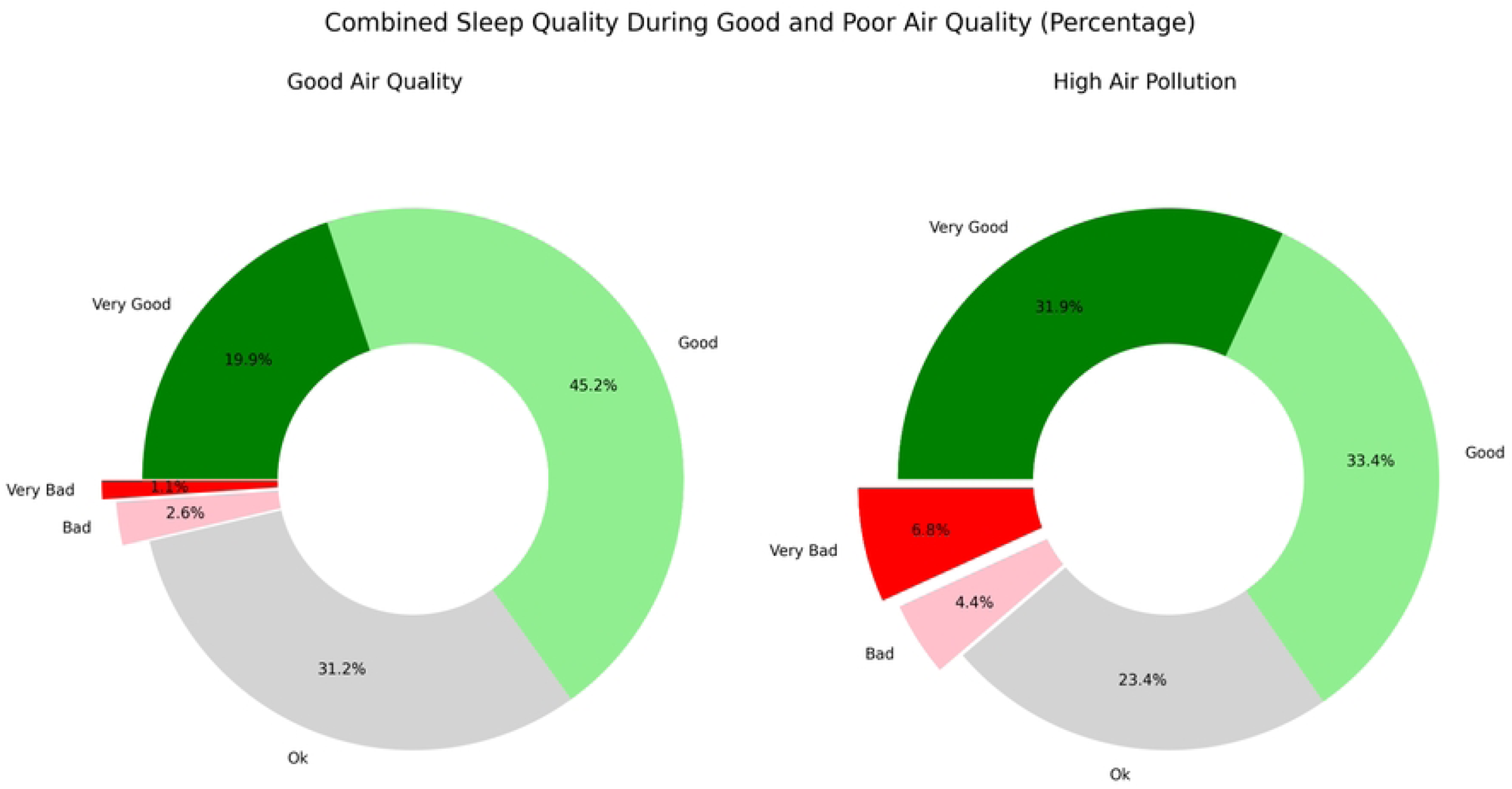
Percentage of reported sleep quality ratings in Delhi and Dhaka combined during good air quality and high air pollution periods. Distribution of self-reported sleep quality during good air quality and high air pollution periods.

For sleep quality, participants living in Delhi reported significantly different outcomes than did those living in Dhaka (p = 0.003). In pooled analyses across Delhi and Dhaka, relative to the 1–4-year reference group, children aged 0 years or 10–14 years were associated with poorer sleep quality (bad or very bad). A lower income (<$100 or $100–$499) was also linked to poorer sleep quality than higher income. Both female (p = 0.040) and male participants (p = 0.032) reported higher sleep quality than did those who preferred not to state their gender.

For general feelings, being aged 15–19 (p < 0.001), 20–24 (p =0.007), or 25–34 (p = 0.018) was associated with more positive feelings (good and very good) than being aged 55 or older.

Compared with the reference group (1–4 years), children aged 0 years, 10–14 years, or 15–19 years were associated with fewer positive feelings (bad and very bad). Compared with the reference group, the female participants reported more positive feelings (p = 0.004), whereas the effect for the male participants was not statistically significant (p = 0.240). Higher income was also associated with more positive feelings, particularly among those earning $1500-$4000 (p < .001) or >$4000 (p < 0.001). The full estimates are provided in S2 Table.

### Health Symptoms

The participants’ self-reported health symptoms varied significantly by air quality. In Delhi, reports of itchy eyes increased from 34.3% (n = 57) during good air quality to 69.3% (n = 138) during high air pollution (p < 0.001). Skin irritation or rash rose from 45.2% (n = 75) to 63.3% (n = 126, p = 0.001), and diarrhea or vomiting rose from 28.3% (n = 47) to 67.8% (n = 135, p < 0.001). Respiratory difficulties also increased from 44.0% (n = 73) to 62.8% (n = 125, p < 0.001), whereas emotional symptoms increased, with low moods increasing from 58.4% (n = 97) to 71.9% (n = 143, p = .007) and anxiety/stress from 62.0% (n = 103) to 72.4% (n = 144, p = .036).

In Dhaka, multiple symptoms were significantly associated with poor air quality. The percentage of itchy eyes rose from 36.8% (n = 39) to 56.6% (n = 194, p < .001). Sore throat increased from 38.7% (n = 41) to 57.1% (n = 196, p =0.001), cough increased from 37.7% (n = 40) to 57.4% (n = 197, p < .001), and headache increased from 28.3% (n = 30) to 62.7% (n = 215, p < .001). Emotional symptoms also increased: low mood from 20.8% (n = 22) to 56.9% (n = 195, p < .001) and anxiety/stress from 22.6% (n = 24) to 53.4% (n = 183, p < .001). Gastrointestinal and skin-related complaints were also greater, with diarrhea or vomiting increasing from 34.0% (n = 36) to 57.1% (n = 196, p < .001) and skin irritation/rash from 34.9% (n = 37) to 55.7% (n = 191, p < .001). Difficulty concentrating also increased from 23.6% (n = 25) to 54.5% (n = 187, p < .001). (See S2 Fig and S3 Table for city-specific details.

When combined across both cities (N = 814), the clearest differences were observed for itchy eyes (35.3% vs. 61.3%, p < .001), diarrhea or vomiting (30.5% vs. 61.1%, p < .001), and respiratory difficulties (36.0% vs. 58.9%, p < .001). Emotional effects were also notable, with low mood (43.8% vs. 62.4%, p < .001) and anxiety/stress (46.7% vs. 60.3%, p < .001) more frequently reported during high air pollution. Headache (48.9% vs. 64.4%, p < .001) and difficulty concentrating (51.8% vs. 61.6%, p = .008) were also significantly more common under high-pollution conditions. Overall, high air pollution was associated with a greater burden of both physical and emotional symptoms (Fig 5; S6 Table).

**Fig 5:**
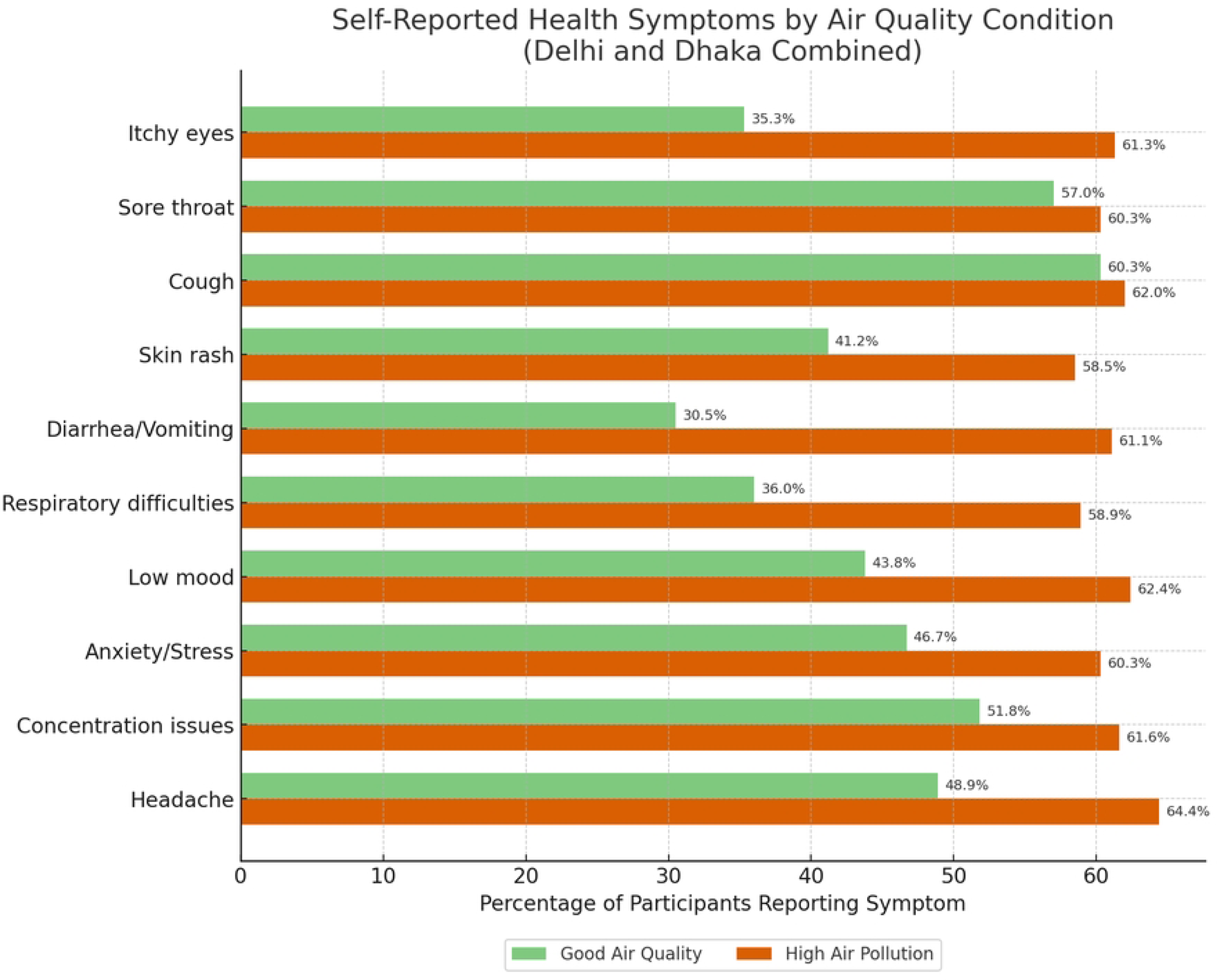
Percentages of participants reporting health symptoms in Delhi and Dhaka combined during good air quality and high air pollution periods. Distribution of self-reported health symptoms during good air quality and high air pollution periods.

Following the chi-square analyses, binary logistic regression was conducted to examine how reported symptoms were associated with air quality periods, city of residence, and participant demographics (Fig 6 and S4 Table).

**Fig 6:**
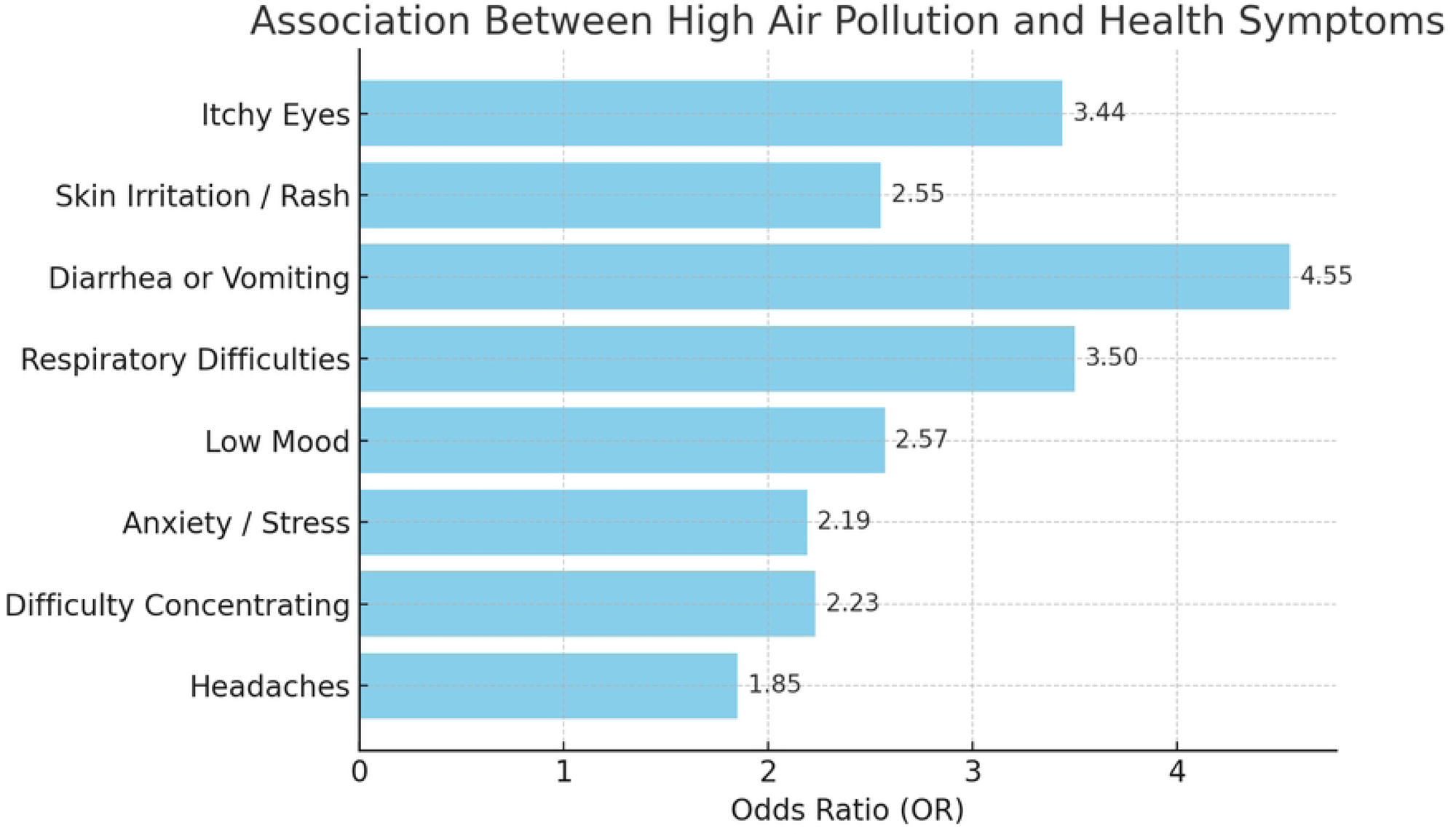
Odds ratios for the associations between high air pollution and self-reported health symptoms. This figure presents adjusted odds ratios (ORs) from binary logistic regression models showing the likelihood of reporting each health symptom during periods of high air pollution compared with good air quality for both cities.

City of residence played a significant role in symptom reporting. The participants in Dhaka consistently reported fewer health issues than did those in Delhi. The odds of reporting symptoms were significantly lower in Dhaka across all measured outcomes. Symptoms such as itchy eyes, skin irritation or rash, diarrhea or vomiting, and headache were moderately reduced (odds ratios (ORs) between 0.50 and 0.54, all p ≤ 0.001). More pronounced reductions were observed for respiratory difficulties (OR = 0.47, p < 0.001), difficulty concentrating (OR = 0.31, p < 0.001), low mood (OR = 0.36, p < 0.001), and anxiety or stress (OR = 0.28, p < 0.001). These patterns suggest a substantially greater health burden in Delhi than in Dhaka, affecting both physical and mental health (S4 Table).

High air pollution periods were also strongly linked to reported health symptom occurrence. Compared with periods of good air quality, periods of high air pollution were associated with significantly increased odds of reporting all health symptoms. The largest effects were observed for gastrointestinal and respiratory symptoms, particularly diarrhea or vomiting (OR = 4.55, *p* < 0.001), respiratory difficulties (OR = 3.50, *p* < 0.001), and itchy eyes (OR = 3.44, *p* < 0.001). Moderate increases were observed for skin irritation or rash (OR = 2.55), low mood (OR = 2.57), anxiety or stress (OR = 2.19), and difficulty concentrating (OR = 2.23), all *p* < 0.001. Headache had a smaller but still significant association (OR = 1.85, *p* = 0.001) (see Fig 6 and S4 Table). Together, these findings highlight the broad impact of high air pollution on both physical and mental health.

Several demographic factors were also associated with symptom patterns. Male participants and those who did not disclose their sex reported significantly lower odds of experiencing physical symptoms (itchy eyes, skin irritation, diarrhoea, respiratory issues) than female participants did. Having a child, especially in the 0-4, 10-14-, or 15-19-years age groups, was linked to higher odds of itchy eyes and mental health symptoms, including low mood, anxiety or stress, and concentration difficulties. Income effects were less consistent, while lower-income groups (<$100 or $100-$499) showed no consistent pattern, participants in the $500-$1499 range had significantly higher odds of reporting skin irritation (OR = 2.97, p = 0.002) and respiratory difficulties (OR = 3.25, p < 0.001). Age also influenced symptom reporting. Compared with parents aged 55 or older, parents aged 25-34 years had substantially greater odds of reporting that their children experienced low mood (OR = 5.67, p = 0.018), anxiety or stress (OR = 7.61, p = 0.009), and difficulty concentrating (OR = 4.64, p = 0.035) (Fig 6 and S4 Table).

High air pollution was a strong and consistent predictor of health symptoms, i.e., both physical and mental symptoms, with the health burden notably greater in Delhi than in Dhaka. Demographic factors, including sex, having a child younger than 18 years, income, and age, further shape individuals’ vulnerability to pollution-related health symptoms.

### Daily activity disruption

Self-reported disruptions to daily activities were significantly more common during high air pollution periods in both Delhi and Dhaka. In Delhi, participants were more likely to report no physical activity during high air pollution (17.6%, n = 35) than during good air quality (8.4%, n = 14). The share of those reporting at least 60 minutes of physical activity fell from 41.6% (n = 69) during good air quality to 17.1% (n = 34) during high air pollution (S5 Table). Similarly, disruptions to daily routines increased under high air pollution: 69.8% (n = 139) reported being late in school or work, whereas 30.7% (n = 51) reported being late during good air quality; 68.8% (n = 137) missed school or work, ranging from 43.4% (n = 72); 66.3% (n = 132) missed a meeting or interview, ranging from 41.6% (n = 69); 63.3% (n = 126) missed healthcare appointments, ranging from 28.3% (n = 47); 65.3% (n = 130) missed social gatherings, ranging from 30.1% (n = 50); and 69.3% (n = 138) needed more family assistance, compared with 44.6% (n = 74) under good air quality (S6 Table).

In Dhaka, 31.5% (n = 108) of the participants reported no physical activity during high air pollution, whereas only 5.7% (n = 6) reported good air quality. Those reporting 60+ minutes of activity decreased from 50.9% (n = 54) to 6.4% (n = 22) (S5 Table). Other disruptions also increased: 60.1% (n = 206) reported being late in school or work (vs. 30.2%, n = 32 during good air), 55.4% (n = 190) missed school or work (vs. 29.2%, n = 31), 55.7% (n = 191) missed a meeting or interview (vs. 22.6%, n = 24), 53.6% (n = 184) missed healthcare appointments (vs. 21.7%, n = 23), 57.4% (n = 197) missed social gatherings (vs. 27.4%, n = 29), and 64.4% (n = 221) needed more family assistance (vs. 26.4%, n = 28) (S6 Table).

Across the combined sample, reports of no physical activity rose from 7.4% (n = 20) during good air quality to 26.4% (n = 143) during high air pollution, whereas reports of 60+ minutes of activity declined from 45.2% (n = 123) to 10.3% (n = 56) (S5 Table). The proportion of participants who were late in school or work rose from 30.5% (n = 83) to 63.7% (n = 345), and the percentage of those who missed school or work increased from 37.9% (n = 103) to 60.3% (n = 327). Similarly, 59.6% (n = 323) missed meetings or interviews during periods of high air pollution, whereas 34.2% (n = 93) missed meetings or interviews during periods of good air quality. Reports of needing more family assistance rose from 37.5% (n = 102) to 66.2% (n = 359). In addition, 58.3% (n = 310) missed healthcare appointments during high-pollution periods compared with 25.7% (n = 70) under good-air quality periods, and 60.3% (n = 327) missed social gatherings (friends/family) compared with 29.0% (n = 79) in good-air quality periods (Fig 7 and S6 Table).

**Fig 7:**
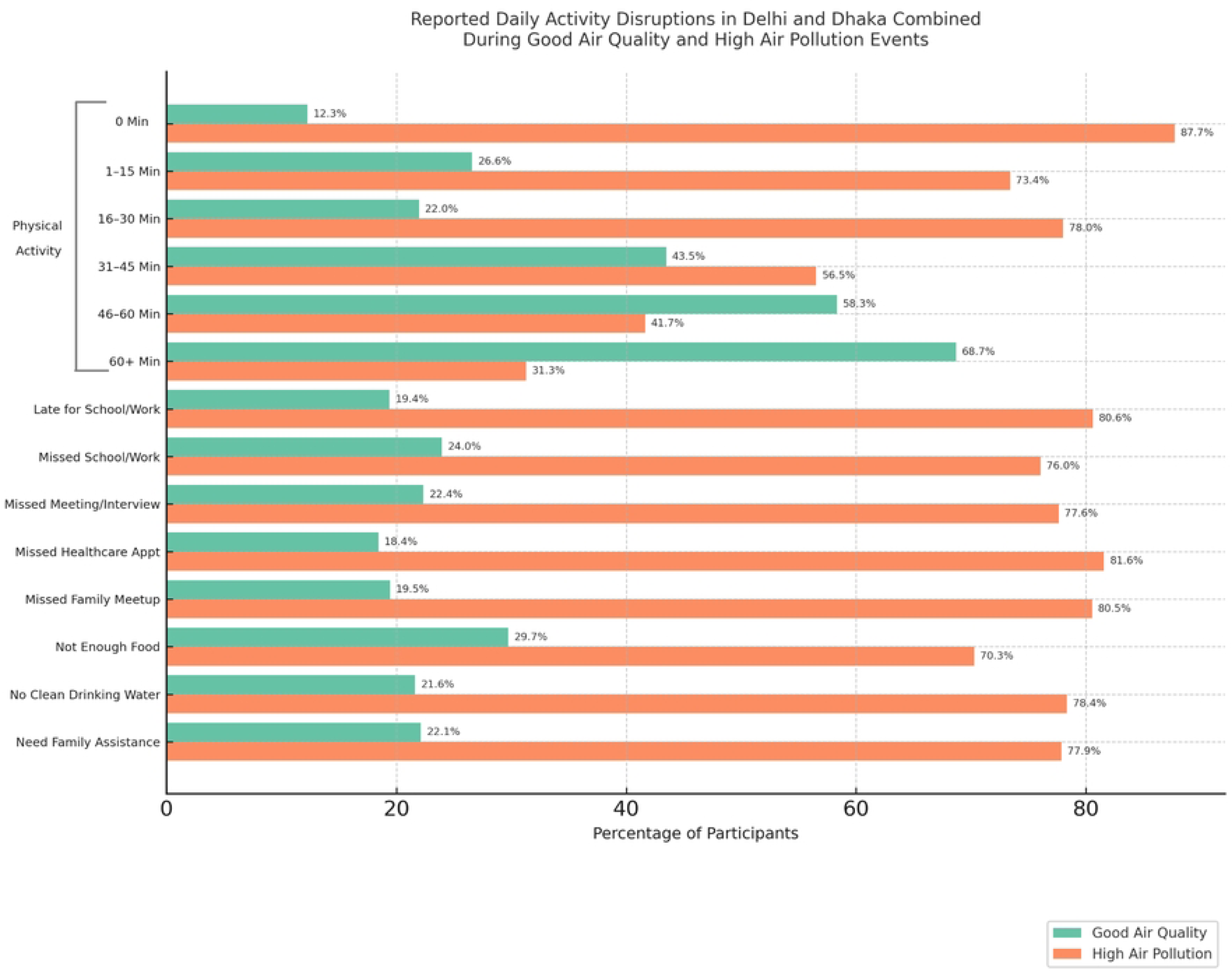
Percentages of participants reporting daily activity disruptions in Delhi and Dhaka combined during good air quality and high air pollution periods. Illustrates the distribution of self-reported disruptions to daily activities during various air quality periods.

All daily activity disruption variables were significantly associated with air quality in the combined sample (all p < 0.001). In both Delhi and Dhaka, participants were more likely to report reduced physical activity; being late or missing school, work, or health appointments; and needing more family assistance during high air pollution (Fig 7). Most disruptions were significant in both cities individually (p < 0.001), although “not having enough food” was significant only in Dhaka (p < 0.001) and in the combined analysis (p = 0.005) but not in Delhi (p = 0.210) (S6 Table). This suggests that air pollution was linked to interruptions in daily routines, with some city-specific variations.

In the chi-square analyses, regression analysis was conducted only for activity variables that were significantly associated with air quality. Ordinal logistic regression revealed that air quality, age, parental status, and income were significantly associated with levels of physical activity (all p < .05). Compared with those in high air pollution conditions, those in good air quality conditions were more likely to engage in more physical activity. In contrast, participants aged 5- 9, 10-14, 15-19, and 20-24 years were less likely to report higher physical activity levels than were those aged 55 years or older. Parents with children aged 0-4, 10-14, or 15-19 years were more likely to report higher levels of physical activity than those without children. Participants with a monthly income of $1500-$4000 had lower odds of higher physical activity than those earning less than $100 (S6 Table).

The participants exposed to high air pollution had more than four times greater odds of being late in school or work (OR = 4.60), missing a meeting or interview (OR = 4.35), missing a healthcare appointment (OR = 4.44), lacking access to clean drinking water (OR = 4.80), and requiring more family assistance (OR = 4.50). Increased odds were also found for missing school or work (OR = 3.76), missing social meetings (OR = 3.95), and not having enough food (OR = 1.88) (Fig 8; S8 Table).

**Fig 8:**
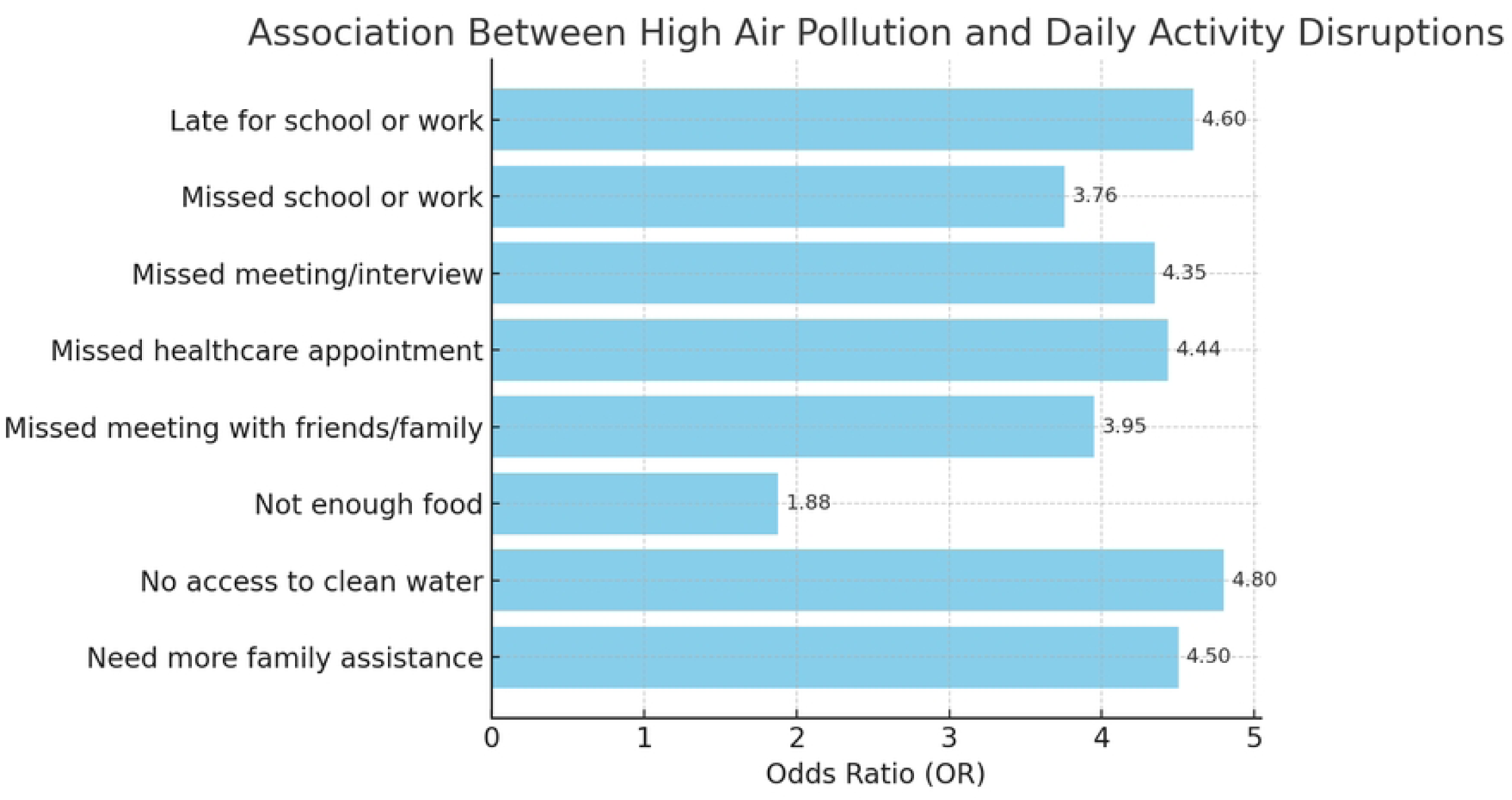
Adjusted odds ratios for the effect of high air pollution on daily activity disruptions. This figure shows the likelihood of experiencing various disruptions during periods of high air pollution compared with good air quality in both cities, based on binary logistic regression models. Odds ratios are adjusted for demographic factors.

### Perceptions and responses to high air pollution

Chi-square analyses revealed significant differences in concern about high air pollution between good- and high-pollution days in both Delhi and Dhaka (p < .001). In Delhi, the highest possible concern level (a score of 10, meaning “extremely concerned about high air pollution”) was much more common on high-pollution days. In Dhaka, participants were also more concerned with high-pollution days than days with good air quality, but instead of most selecting the maximum score, their concern levels were spread across both moderately high (scores 6-8) and very high (scores 9-10) ratings (S3 Fig).

The perceptions of community preparedness and response to high air pollution also varied significantly by air quality in both Delhi and Dhaka (p < .001). In both cities, higher satisfaction levels (“very satisfied” or “somewhat satisfied”) were more common on high-pollution days, although the distributions differed across locations (S9 Table).

These significant differences in concern and satisfaction (S3 Fig and S9 Table) were reflected in the participants’ qualitative responses, which revealed how they envisioned addressing the challenges of high air pollution . Five themes were identified (see S10 Table) for detailed themes and illustrative quotes; see Figure 9 for the distribution of themes and frequencies). The most common approach was Cleanliness & Environment, with repeated calls for cleaner streets and more greenery: *“It would be good if the mud or garbage around the road gets cleaned up”* and *“Plant more trees or reforestation.”* Values & Community highlighted moral and social responsibilities, with participants stressing *“We need God”* and the need to *“Vote great leaders that are accountable.”* In Health & Wellbeing, participants pointed to stronger services and education, for example, *“Free and equipped hospitals”* and *“Health education.”* Pollution & Technology Solutions targeted sources directly, including *“Remove fuel cars,” “Use electric vehicles,”* and *“Ban smoking in public places.”* The other category captured short or unclear remarks, which were retained for completeness. These perspectives suggest that participants’ concerns extended beyond immediate health impacts to broader visions of cleaner environments, stronger communities, improved services, and better governance.

**Fig 9:**
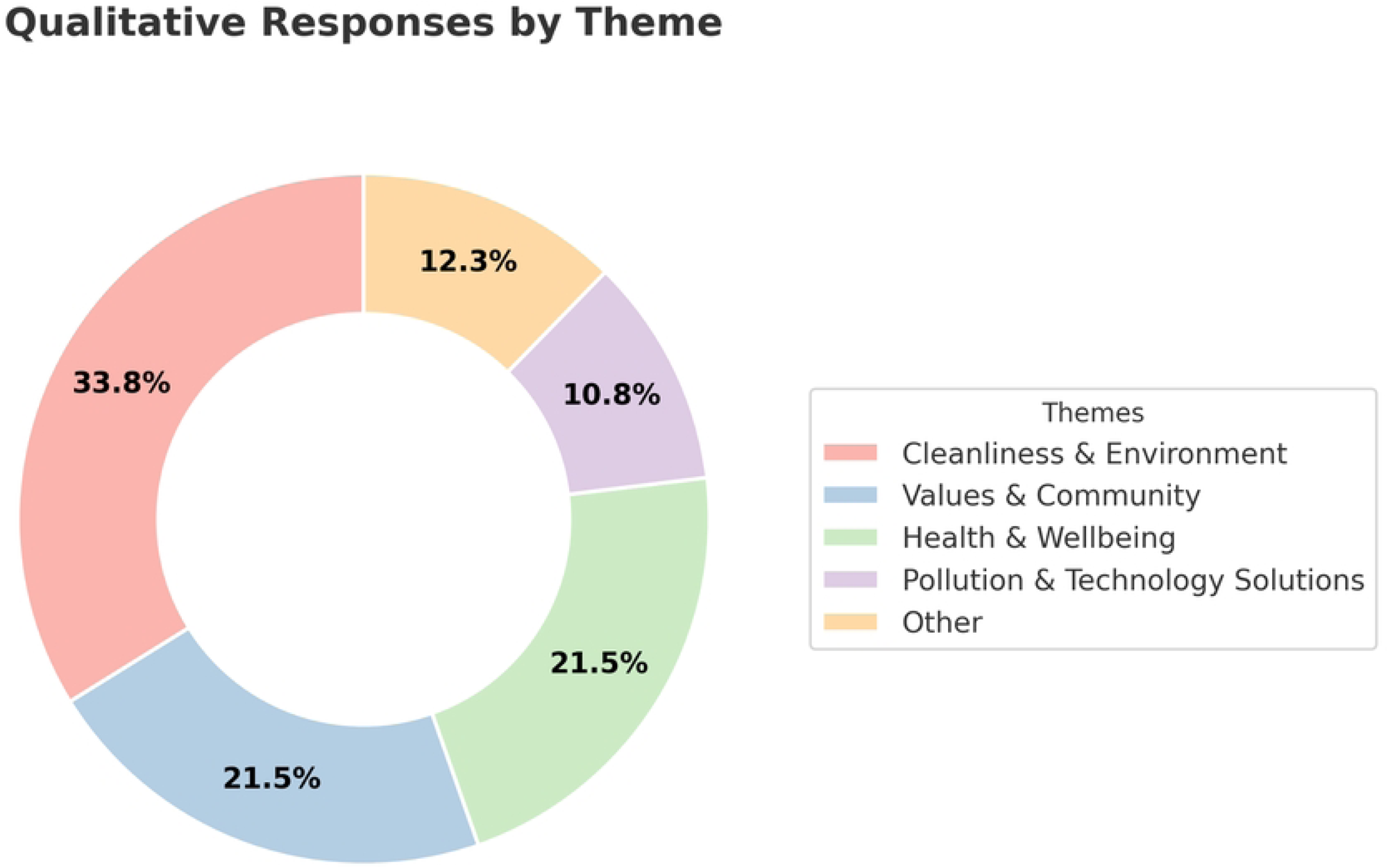
Distribution of qualitative responses across five themes. Donut chart showing the percentage of responses categorized into the following categories: cleanliness & environment, values & community, health & wellbeing, pollution & technology solutions, and other. The percentages represent the share of total responses (n = 145).

## Discussion

This study compared periods of good air quality and high air pollution in Delhi and Dhaka, identifying consistent differences in self-reported well-being, health symptoms, and daily activities (Figs. 3-7; S2 Table, S3 Table, and S8 Table). A mixed-methods approach was used, combining quantitative analyses (descriptive statistics and regression models adjusted for demographic differences) with qualitative thematic analysis of open-ended responses in NVivo.

### Health, well-being, and daily activity disruption

Self-reported well-being, including both general feelings and sleep quality, was significantly associated with air quality conditions [1,11,13]. Participants reported more negative feelings and poorer sleep during high-pollution periods, consistent with existing evidence linking air pollution to disruptions in circadian rhythms, increased psychological distress, and reduced sleep efficiency [6,11]. These effects are thought to arise through pathways such as systemic inflammation, oxidative stress, and heightened autonomic arousal, which can interfere with emotional regulation and restorative sleep [6,30]. Younger participants and those from lower-income brackets appeared more vulnerable, potentially reflecting greater psychosocial stress and fewer protective resources [2,14]. While regression models did not show air quality as a statistically significant predictor after adjustment, the observed patterns suggest that ambient pollution may still influence subjective well-being through indirect or cumulative effects that merit further longitudinal investigation [1,11,13].

After adjusting for demographic characteristics, high air pollution remained strongly linked to increased odds of reporting itchy eyes, skin irritation or rash, diarrhea or vomiting, respiratory difficulties, low mood, anxiety or stress, difficulty concentrating, and headaches (Figs. 5-6; S4 Table and S8 Table). These findings are consistent with evidence that pollutants such as particulate matter can affect both physical and mental health through airway inflammation, oxidative stress, and effects on the nervous system. For example, ocular irritation may result from pollutant-induced inflammation of the eye surface, leading to conditions such as conjunctivitis and irritation [9]. Skin problems, including dermatitis and hyperpigmentation, have been linked to similar inflammatory and oxidative pathways [30]. Respiratory and neurological effects may arise when systemic inflammation and oxidative stress damage the blood‒brain barrier or cause direct injury to nerve tissue [6,31]. Emotional and cognitive impacts, such as low mood, anxiety, and reduced concentration, are in line with studies showing that air pollution can increase internalizing symptoms, alter brain structure and function, and increase the expression of markers of neuroinflammation [11].

When adjusted for demographic factors, younger participants were more likely to report stress, low mood, and concentration problems, potentially reflecting heightened psychosocial and neurological sensitivity to air pollution. This aligns with evidence linking pollution exposure to oxidative stress, inflammation, and neuropsychiatric disorders, processes that may disproportionately affect younger populations during key life transitions [6,11,31]. Income effects were less consistent: middle-income groups reported fewer physical symptoms, such as irritation, but had higher odds of respiratory difficulties. These patterns may reflect differences in housing quality, occupational exposure, or healthcare access across socioeconomic strata, which is consistent with research showing that socioeconomic position affects long-term pollution-related health risks and mortality outcomes [2,32,33]. These associations remained even after adjusting for demographic factors, indicating that the health impacts were not only due to those who were exposed to high air pollution. However, factors such as gender, parental status, and income still influence the likelihood of reporting certain symptoms [2,34].

The participants exposed to high air pollution were more likely to be late in school or work, miss school or work, miss important meetings or healthcare appointments, lack access to clean drinking water, and require more family assistance. In several cases, the adjusted odds exceeded four, indicating a substantial impact of pollution episodes on everyday functioning (Figs. 7-8; S6 Table and S8 Table). These findings are consistent with prior evidence. School absences increase with increasing air pollution [35,36]. Similarly, workplace studies have shown lower productivity and higher absenteeism under polluted conditions, even in indoor and service-sector settings [13,37]. The reduction in physical activity is consistent with evidence that individuals limit outdoor exercise during high air pollution, both through avoidance behaviour and symptom-driven constraints [37]. Missed healthcare appointments may reflect transport and service barriers, a well-documented driver of no-shows [38,39], and have also been observed during wildfire smoke episodes when mobility and clinic operations are disrupted [40].

City-specific differences highlight structural vulnerabilities. In Dhaka, participants reported food shortages and water access problems during high air pollution, which are patterns not observed in Delhi. Dhaka’s unplanned urbanization and fragile water, sanitation, and hygiene (WASH) infrastructure heightens the risk of service interruptions during pollution episodes, a period when absenteeism and low productivity are also more common [4,13,37,41]. Haze-related transport slowdowns and market disruptions have been shown to affect food distribution and pricing [10,42], likely explaining the Dhaka-specific food effects observed in this study. Other studies have revealed more complex patterns. High air pollution has been associated with increases in outpatient visits for acute conditions such as respiratory and cardiovascular disease [43,44], suggesting that urgent care may increase even as routine care is missed. Similarly, some populations shift activities indoors when facilities are available [45]. These differences highlight how resources and infrastructure shape behavioural responses to high air pollution.

After adjusting for demographics, high air pollution remained an independent predictor of reduced physical activity. Younger participants (aged 5-24) were less active, likely reflecting parental restrictions, whereas parents of young or school-aged children reported more activity due to caregiving responsibilities [46]. Those with incomes in the $1,500–$4,000 income bracket were less active than those in the lowest-income group were, which is consistent with evidence that higher-income individuals generally engage in more sedentary behavior and less regular activity despite occasional intense exercise [33], whereas low-income individuals often accrue activity through physically demanding work or active transportation [32].

Overall, this study suggests that air pollution disrupts not only health but also daily activities such as education, employment, mobility, and access to basic services. These effects remained after accounting for demographic factors, showing that they are not explained by population composition alone. Instead, they result from behavioural avoidance, pressure on infrastructure, and economic disruption, with especially strong effects in lower-resource urban settings. Such disruptions represent hidden social and economic costs of air pollution that extend beyond its direct health impacts.

### Public Perceptions and Policy Implications

Public concern was greater during high-pollution periods (S9 Table). The qualitative responses emphasized interventions such as tree planting, waste management, cleaner transport, and accountable leadership (Fig 9 and S10 Table). These priorities closely align with WHO recommendations for urban air quality management and with regional analyses that advocate multisectoral strategies and low-emission transport options [1,47]. The overlap between community priorities and expert guidance indicates the potential for public engagement in designing and implementing air quality policies.

### Limitations and Future Directions

This study has several limitations that are important to consider. Since the findings are based on self-report surveys, they may reflect how participants remembered or chose to describe their experiences, which introduces the risk of bias. Cultural and linguistic differences may also have shaped how symptoms and feelings were expressed, even though translations were carefully reviewed. The cross-sectional design offers valuable insights into the associations between air quality and reported experiences; however, it cannot demonstrate whether one factor directly affects another.

Air quality was measured via city-wide monitoring data rather than personal exposure assessments, which means that the reported levels may not fully capture the variation in what each participant experienced in the changes in air quality. Recruitment through online advertisements allowed the study to reach young people quickly across different settings; however, it excluded those without internet access and may not represent the full diversity of the population.

In addition, differences in participation between periods may reflect non-equivalent sampling windows (weekday/weekend and holidays). In Delhi, the high-pollution window (Thursday–Friday, 9–10 January) fell on routine school/work days, whereas the good-air window (Saturday, 15 March) followed Holi but was not a routine school holiday; participation opportunities were therefore likely similar across periods. In Dhaka, the high-pollution window (Tuesday–Wednesday, 21–22 January) comprised school/work days, while the good-air window (Thursday–Sunday, 13–16 March) overlapped the Friday–Saturday weekend, when many students are at home. These calendar effects may partly explain the differences in numbers between periods and could contribute to differential availability and non-response. Finally, while self-reported health and activity outcomes provide useful perspectives, the absence of objective measures such as medical records or wearable devices limits the accuracy of the results.

Future research can build on this study in several ways. Personal exposure monitoring would provide more precise insights into how individuals are affected, moving beyond city-wide estimates. The incorporation of objective measures, such as medical records, wearable activity trackers, or sleep monitors, would also strengthen the evidence base. Longitudinal designs could help identify how repeated or long-term exposure influences health and daily life, going beyond the snapshot provided here. Intervention studies are another important next step, testing practical approaches such as air filtration, mask use, or adjustments to school and work routines. Finally, expanding research to additional cities across South Asia and beyond would allow for comparisons across different contexts, helping to identify both common patterns and location-specific needs.

## Conclusion

The convergence of physical, emotional, and behavioural impacts observed here underscores the multifaceted burden of high air pollution in urban South Asia. The alignment between community-identified solutions and established public health recommendations presents an opportunity for participatory, evidence-based interventions that address both the causes and the consequences of high air pollution.

## Declarations

### Ethics approval and consent to participate

This study was approved by the London School of Hygiene & Tropical Medicine Research Ethics Committee (Reference 31044). All participants provided informed consent prior to completing the survey. For participants under 18 years of age, parents or legal guardians provided consent and completed the survey on behalf of their child. The offices of the mayors in Delhi and Dhaka were also informed of the study prior to data collection.

### Consent for publication

Not applicable. This manuscript does not contain any person’s data in any form (including images, videos, or identifiable details).

### Availability of data and materials

The datasets generated and analyzed during the current study are openly available in the LSHTM Data Compass repository, https://doi.org/10.17037/DATA.00004813. All additional information and summary data supporting the findings are included in this article and its supplementary information.

## Authors’ contributions

Constance Bwire led the conceptualization and methodology, conducted the investigation, curated and analyzed the data, prepared visualizations, managed the project, and drafted the original manuscript. Robert Hughes contributed to conceptualization and methodology, provided supervision, secured funding, and revised the manuscript. Rachel Juel contributed to conceptualization and methodology. Gabrielle Bonnet, Ana Bonell, James Milner, and Shunmay Yeung contributed to manuscript review and editing. All authors read and approved the final version.

## Data Availability

All relevant data are openly available through the LSHTM Data Compass repository at https://doi.org/10.17037/DATA.00004813. Additional summary data and supplementary materials are provided within the manuscript and its supporting files.

## Acknowledgements

The authors gratefully acknowledge the children, young people, and parents who generously shared their time by participating in the real-time survey activities. Appreciation is also extended to local teams and partner organizations in Delhi and Dhaka for their support in coordinating and implementing data collection. In particular, we would like to thank the Empower Agency for their assistance with participant recruitment and for providing training to staff engaged in online data collection. The authors also thank the wider CCC Action Lab team for their collaboration throughout the study.

## Supplementary Information

**S1 Text (DOCX): Protocol for identifying CCC Action Lab focal cities.** Criteria and procedures for selecting the 178 cities included as focal sites for air pollution period identification in this study.

**S1 File (DOCX): Survey recruitment and digital advertisement strategies.** Examples of the digital advertisements used to recruit participants in Delhi and Dhaka via Meta and Google platforms.

**S2 Text (DOCX): Data collection instrument (Hindi).** Full survey questionnaire in Hindi, as used in Delhi.

**S3 Text (DOCX): Data collection instrument (Bengali).** Full survey questionnaire in Bengali, as used in Dhaka.

**S4 Text (DOCX): Sample mayor’s email.** Example of the outreach email used to inform mayors’ offices in Delhi and Dhaka about the study prior to data collection.

**S1 Table (XLSX): Demographic characteristics.** Breakdown of age, gender, parental status, and income for survey participants.

**S1 Fig (TIFF): General feelings.** Figure showing the percentage of participants reporting general feelings during good air quality and high air pollution periods per city.

**S2 Table (XLSX): Well-being (general feelings and sleep quality).** Participant-reported measures of well-being and sleep quality by air quality period.

**S2 Fig (TIFF): Health symptoms.** Figure showing the percentage of participants reporting physical and emotional health symptoms during good and high air pollution periods per city.

**S3 Table (XLSX): Self-reported health symptoms.** Data summarizing physical and emotional health symptoms reported across air quality periods.

**S4 Table (DOCX): Logistic regression results for health symptoms.** Adjusted odds ratios for the likelihood of experiencing symptoms during high-pollution compared with good air quality periods.

**S5 Table (XLSX): Physical activity.** Self-reported levels and changes in physical activity during good and high air pollution periods.

**S6 Table (DOCX): Ordinal logistic regression for physical activity.** Adjusted regression models assessing associations between air quality, demographics, and levels of physical activity.

**S7 Table (XLSX): Disruptions in daily activities.** Interruptions to school, work, healthcare, and caregiving activities reported during high-pollution periods.

**S8 Table (DOCX): Logistic regression results for daily activity disruptions.** Adjusted odds ratios for the likelihood of experiencing daily activity disruptions during high-pollution periods.

**S3 Fig (TIFF): Concerns about air pollution.** Figure showing the distribution of participant concern levels in Delhi and Dhaka across air quality periods.

**S9 Table (XLSX): Concern and satisfaction distributions.** Tabulated data on concern levels about high air pollution and satisfaction with city responses.

**S10 Table (TIFF): Community preparedness and response.** Figure showing participant satisfaction with city preparedness and response to high air pollution in Delhi and Dhaka.

**S10 Table (XLSX): Thematic analysis of open-ended responses.** Qualitative codebook, themes, and illustrative quotes on participants’ suggestions for improving urban sustainability.

